# Probability and Estimated Risk of SARS-CoV-2 Transmission in the Air Travel System: *A Systemic Review and Meta-Analysis*

**DOI:** 10.1101/2021.04.08.21255171

**Authors:** Jenna K. Pang, Stephen P. Jones, Lindsay L. Waite, Nels A. Olson, Robert J. Atmur, Joshua J. Cummins

## Abstract

As an emerging virus, SARS-CoV-2 and the risk of transmission during air travel is of high interest. This paper estimates the probability of an infectious index passenger in the air travel system transmitting the SARS-CoV-2 virus to a fellow passenger during air travel. Literature was reviewed from May–September 2020 to identify COVID-19 cases related to the air travel system. The studies were limited to publicly available literature for passengers starting in January 2020; studies on other persons such as flight crews were not reviewed. A novel quantitative approach was developed to estimate air travel transmission risk that considers secondary cases, the overall air travel passenger population, and two correction factors for asymptomatic transmission and underreporting. There were at least 2866 index infectious passengers documented to have passed through the air travel system in a 1.4 billion passenger population. With correction factors, the global risk of transmission during air travel is 1:1.7 million. Uncertainty in the correction factors and a 95% credible interval indicate risk ranges from 1 case for every 712,000 travelers to 1 case for every 8 million travelers. The risk of COVID-19 transmission on an aircraft is low, even with infectious persons onboard.

## 1. Introduction

In late 2019, an outbreak of a novel coronavirus disease, now known to be caused by the severe acute respiratory syndrome coronavirus 2 (SARS-CoV-2) virus and named coronavirus Disease 2019 (COVID-19), was first documented in Wuhan, China [1]. By January 2020, the disease was confirmed to have spread to other parts of Asia [2], and by February 2020 cases were being seen around the globe [3]. The World Health Organization (WHO) declared the global outbreak a pandemic on March 11, 2020 [1]. As the pandemic continued, the air travel industry was severely impacted with a nearly 95% drop from 2019 passenger levels in a matter of weeks [4]. With segments of the global population continuing to travel, however, questions emerged regarding the risk of disease transmission in the air travel system, and specifically in the aircraft cabin. Although the risk of SARS-CoV-2 transmission among the flying public is perceived to be high, a data-driven approach is required to determine the actual range of risk.

It is important to differentiate between COVID-19 cases imported by air travel (i.e. an index passenger travels to a new location and transmission occurs on the ground) and cases as the result of transmission during air travel (i.e. in the airplane cabin); the risk of infection via air travel transmission is not equal to the probability of an infectious person aboard an airplane. This paper quantifies the risk of SARS-CoV-2 transmission during air travel by estimating the risk of transmission in the air travel system from data collected during the pandemic and corrects for known data limitations. Acknowledging that COVID-19 testing and reporting requirements vary by locality and can be difficult to connect with air travel, uncertainties are included in these risk calculations to more conservatively estimate risk. This paper is limited to publicly available literature which primarily considers infectious passengers and potential secondary transmissions during air travel; transmission risk among airport personnel such as Transportation Security Administration (TSA) agents or crew-to-crew transmission are not addressed.

Research on disease transmission in flight from other viruses such as from SARS-CoV-1 [5], Influenza [6], and Tuberculosis [7] has shown that the risk of known communicable diseases being transmitted aboard aircraft is low [8]. Generally, investigations have found low SARS-CoV-2 transmission rates, such as was the case for the well-documented Milan-South Korea repatriation flights in March 2020 (one secondary case on each flight) where passengers wore masks and participated in mandatory screening and quarantine [9] and the Singapore-Hangzhou flight in early January 2020 with fifteen index passengers but only one potential secondary case [10]. A notable exception is the VN54 flight between London and Hanoi on March 1st-2nd in which fifteen secondary cases were presumed to be related to one index passenger [11]. However, the VN54 flight study’s non-quantitative analysis and the large number of days between flight and COVID-19 testing of passengers brings into question if each of these fifteen COVID-19 cases was the result of transmission in the airplane cabin or exposures after the flight from other social interactions.

## 2. Methods

### 2.1. Literature Review for Air Travel Transmission Cases

A literature review was performed from May to September 2020 to identify index and secondary COVID-19 cases related to transmission in the air travel system for travel from January to September 2020. Index cases are those who tested positive for COVID-19 upon arrival at their destination. Thus, based on the known incubation period of the disease, these passengers were identified as infected prior to entering the air travel system. Secondary cases are persons who travelled on a flight with an index passenger, tested positive for COVID-19 after the flight, and were identified through contact tracing to have most likely been infected via transmission from the index passenger. Publications were collected using English-language search terms linking COVID-19 and air travel from two primary paths: review of daily news articles and social media posts (referencing published research) or direct search of scientific publication repositories (including arXiv.org, Clarivate, LitCovid, and the CDC’s COVID-19 research database). Additional papers were identified by biomedical experts at International Air Transport Association (IATA) and the Commonwealth Scientific and Industrial Research Organization (CSIRO). As a quality measure, only studies accepted by a journal for publication or by a preprint server were reviewed due to the expected higher quality of information on flights taken by an index passenger and subsequent contact tracing. Two additional studies performed by IATA and the United States Centers for Disease Control and Prevention (CDC) were added to the review due to organizational oversight and data collection capabilities, despite the authors’ limited visibility into the data collection methods and sources. Acknowledging that the variability of recommended behaviors and policy across airlines and governments during the pandemic likely leads to confounding issues in all papers in this study (i.e. mask use, seating), as well as a lack of similar study design, no results were disqualified in the interest of making a conservative, worst case estimate of risk.

The majority of publicly available studies focused on infectious passengers and related transmissions, therefore the study was limited to flights with passenger index cases and did not include transmissions amongst air crew, ground crews, or airport staff. Multiple experts reviewed each study and independently extracted flight information (date, destination and arrival cities, and time of flight), number of index cases, and number of secondary cases as well as available data on passenger behavior. Conservatively, all probable secondary cases identified in the literature are included in risk calculations. Study duplications were recognized by comparing flight information and the study with more information about passenger contact tracing was kept in the review. Discussion was held among reviewers to confirm validity of identified secondary cases being infected in the air travel system, most specifically the airplane cabin, by studying given information about passenger behavior and comparing the index infectiousness period, symptom onset, and positive test dates to COVID-19 incubation period known in September 2020. Group consensus on the number of index and secondary cases was required before proceeding on the study. To verify reasonable interpretations of the review, conversations were held with IATA and the organization’s Medical Advisor, Dr. David Powell (author of [29]), to ensure all publically known flights with transmission were included in the review and to consider the organization’s understanding of air travel risk.

The majority of studies in the literature review focused on a single flight with known secondary cases. Other studies outlined multiple flights, but made qualitative statements on risk based only on the select handful of flights reviewed. News stories released by media and not included in this review indicated index cases aboard many more flights than covered in the published papers, but did not follow up on contact tracing results or identification of secondary cases. A nuanced understanding of air travel risk should therefore consider the full range of air travel experiences during the pandemic from a flight with multiple secondary cases and no mitigation techniques to a flight with no infectious passengers.

### 2.2. Calculation of Estimated Risk of Transmission

To estimate the risk of air travel transmission, a quantitative approach was taken that considers secondary cases and the overall air travel passenger population. A novel approach to transmission risk was developed which uses two correction factors for asymptomatic transmission and underreporting to address the confounding factors and study heterogeneity identified during the literature review and to more conservatively estimate risk. The number of infected persons in the transmission events identified in Section 2.1 was determined through either mandatory screening or self-reporting, and it is likely that this count of infected persons is much lower than the true number of persons who have been infected while flying. Therefore, a number of caveats must be considered for potential error in the risk calculation. The first is the number of people who are infected, but are asymptomatic. Per the Centers for Disease Control and Prevention (CDC), asymptomatic infections are nominally estimated to be 40% of the total infections and asymptomatic individuals are estimated to be 75% as infectious as a symptomatic individual [12]. This indicates a factor of 1.3x if assuming asymptomatic passengers are not fully counted as index or secondary cases in the data. The second issue is the number of unreported COVID-19 cases, both index and secondary, in relation to air travel which includes:

- Symptomatic passengers who seek medical attention for symptoms, but do not inform medical personnel of their travel;
- Symptomatic passengers who seek medical attention for symptoms, but medical personnel fail to report travel; and
- Symptomatic passengers who do not seek medical attention.

To account for underreported air travel cases, the number of unreported COVID-19 cases in the general global population is assumed to apply. Recent estimates indicate global under-reporting to be a 10-fold [13] [14], 23-fold [15] and 54-fold [16] multiplier of the current reported cases. With these caveats, the equation for estimated risk of transmission is as follows:

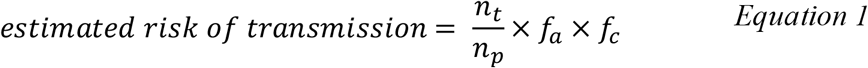

where *n*_*t*_ is the number of secondary cases due to air travel transmission, *n*_*p*_ is the total passenger population, *f*_*a*_ is the factor to account for number of and infectiousness of asymptomatic cases, and *f*_*c*_ is the factor to account for number of unreported cases.

## 3. Results

### 3.1. COVID-19 Transmission Events in the Air Travel System

During the pandemic it has been estimated that, even with a significant decline in air travel, approximately 1.4 billion passengers traveled by air between January and September 2020 [17]. As of August 2020, there were at least 2866 index passengers documented to have passed through the air travel system (Table 1). Due to the widespread prevalence of the disease and presence of pre-symptomatic, asymptomatic, and mildly symptomatic cases in the population, it is known that there necessarily have been additional COVID-19 infectious passengers entering the travel system, beyond the identified index cases. Nonetheless, review of publicly available data reveals fewer than 50 documented potential secondary cases associated with air travel during the pandemic. Mask use on reviewed flights ranged from use unknown to mandatory N95 use.

**Table 1.** Flight, passenger and reference list for index persons and secondary cases due to transmission.

| Flight Information |  | Affected Persons |  | Author |
| --- | --- | --- | --- | --- |
| Date | Country/City Pair | Index | Secondary (due to Transmission) |  |
| General IATA Data |  | 1100 | 3 | IATA [33] |
| General CDC Data |  | 1600 | 0 | Duncan [34] |
| 1/22/2020 | China-Toronto | 1 | 0 | Schwartz et al. [24] |
| 1/24/2020 | Singapore-Hangzhou | 15 | 1 | Chen et al. [10] |
| 2/20/2020 | Tokyo-Tel Aviv | 2 | 0 | Freedman and Wilder-Smith [19] |
| 2/24/2020 | Bangui (CAR)-Paris | 1 | 1 | Eldin et al. [25] |
| 2/26/2020 | Multiple-Greece | 21 | 5 | Pavli et al. [20] |
| 3/2/2020 | London-Hanoi | 1 | 15 | Khanh et al. [11] |
| 3/9/2020 | Tel Aviv-Frankfurt | 7 | 2 | Hoehl et al. [26] |
| 3/10/2020 | Boston-Hong Kong | 2 | 2 | Choi et al. [27] |
| 3/11/2020 | New York City-Taipei | 11 | 0 | Freedman and Wilder-Smith |
| 3/19/2020 | Sydney-Perth | 11 | 11 | Speake et al. [28] |
| 3/31/2020 | Milan-South Korea | 6 | 1 | Bae et al. [9] |
| 4/3/2020 | Milan-South Korea | 3 | 1 | Bae et al. |
| 6/20/2020 | Dubai-Hong Kong | 85 | 2 | Freedman and Wilder-Smith; IATA [29] |
| **Total** |  | 2866 | 44 |  |

Table 1 documents the index and secondary cases for passengers who have flown during the pandemic. International Air Transport Association (IATA) and CDC data are included for thoroughness with the acknowledgement that limited details are known about the data methods and sources and that case counts may be duplicates of other reported cases.

For the purpose of this study, transmission from the index passenger was conservatively assumed to have occurred inside the aircraft cabin in each of these documented cases, although transmission could have occurred elsewhere, such as in the airport. For example, the 15 secondary cases associated with the London-Hanoi flight on March 2, 2020 may not all be the result of in-flight transmission. All of these 15 passengers had either hotel, cruise ship, secondary transportation, travel to a second country, or known contact with other infectious persons not from the flight, or combinations thereof, after the flight from London to Hanoi and prior to testing [11]. Based on the arrival date of the flight (March 2) and the earliest date of the subsequent reverse transcriptase polymerase chain reaction (RT-PCR) tests (March 6), it is possible that the potential secondary cases were infected post-flight since a positive test via PCR can occur on average 2 to 5 days after infection [18]. The testing of the London to Hanoi passengers did not include genomic analysis of the virus and therefore it was not possible to demonstrate that all of the secondary cases map to the same index case by sequence similarity.

For these reasons, the causal connection between the London to Hanoi flight and the eventual positive test results for the 15 cases is in question.

### 3.2. Epidemiological Study of SARS-CoV-2 in Flight

In comparison to the contact tracing performed for the London-Hanoi flight in Section 3.1, other studies were able to more clearly trace index and secondary case relationships to a flight. An epidemiological study of the January 24, 2020 flight from Singapore to Hangzhou, China with a number of confirmed COVID-19 cases on board (Figure 1) was recently completed [10]. This flight, which lasted approximately five hours, was loaded at 89% capacity. Upon arrival at the destination, all passengers and crew were quarantined and tested as necessary. The study determined that of the 16 passengers who ultimately tested positive for COVID-19, 15 of the passengers were index cases on the flight due to the timing of symptom onset; if they had been infected on the flight, they would not have had symptoms immediately after the flight. The index cases had membership in one of three tour groups with other passengers who also tested positive for COVID-19 and originated in Wuhan. Only one passenger was identified to be a potential secondary case of SARS-CoV-2 in flight due to a different city of origin (Hangzhou), membership in a different tour group with no recorded cases of COVID-19, and reported activity of temporarily moving to an unassigned seat surrounded by infectious and symptomatic passengers during the flight as indicated in Figure 1. All 16 passengers wore masks during air travel, but removed their masks during meal time and to drink water; the mask of the secondary case was not worn tightly and did not cover their nose when sitting in the unassigned seat.

**Figure 1:**
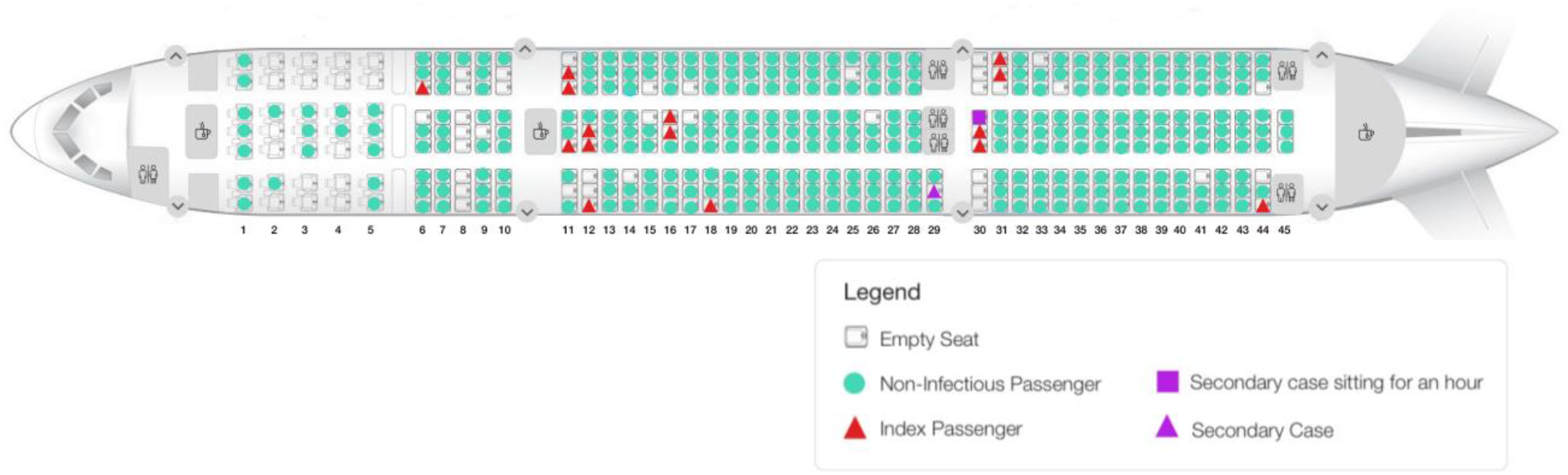
Seat map of flight with 15 index passengers and one possible secondary case.

### 3.3. Estimated Risk of Transmission

The secondary cases listed in Table 1 are for over 10 flights that occurred March 2020 or earlier, while flight traffic was still high, in areas like Europe and the United States. To take a conservative position on risk, transmission risk was calculated for three time periods of interest: (1) January–June 2020 during which the transmission events listed in Table 1 occurred, (2) the month of March 2020 when the global spread of COVID-19 was occurring, and (3) April– September 2020 to account for the sharp drop in worldwide air travel and increased use of COVID-19 testing. For a slightly more conservative risk calculation, the 3 secondary cases recorded by IATA were included in the multi-month risk calculations (January-June 2020, April-

September 2020) based on the assumption that March was a well-documented month, so it is unlikely that these cases are duplicates of the transmission events in that month. Passenger counts were derived from International Civil Aviation Organization (ICAO) monthly reporting of actual passenger totals [17]. Table 2 lists the estimated risk of SARS-CoV-2 transmission in the air travel system for the global public as calculated with Equation 1.

**Table 2.** Estimated risk of SARS-CoV-2 transmission in the air travel system for the global public in 2020.

| Dates | $n_t$ | $n_p$<br>(million) | $f_a$ | $f_c$ | Risk | |
| --- | --- | --- | --- | --- | --- | --- |
|  |  |  |  |  | Numerical | 1 in X |
| January-June 2020 | 44 | 965 | 1.3 | 10 | $5.927 \times 10^{-7}$ | 1,687,063 |
| | | | | 23 | $1.363 \times 10^{-6}$ | 733,506 |
| | | | | 54 | $3.201 \times 10^{-6}$ | 312,419 |
| March 2020 | 31 | 171.3 | 1.3 | 10 | $2.353 \times 10^{-6}$ | 425,062 |
| | | | | 23 | $5.411 \times 10^{-6}$ | 184,810 |
| | | | | 54 | $1.270 \times 10^{-5}$ | 78,715 |
| April-September 2020 | 6 | 552 | 1.3 | 10 | $1.413 \times 10^{-7}$ | 7,076,923 |
| | | | | 23 | $3.250 \times 10^{-7}$ | 3,076,923 |
| | | | | 54 | $7.630 \times 10^{-7}$ | 1,310,541 |

For 44 secondary cases from January–June 2020 with a 1.3-fold factor for asymptomatic persons (*f*_*a*_) and a 10-fold factor for underreporting (*f*_*c*_) against a population of 968 million travelers, the risk of being infected with SARS-CoV-2 in an airplane cabin is estimated to be 5.927×10^−7^ or 1:1.7 million. A 54-fold multiplier of reported cases gives a risk of 3.201×10^−6^ or a 1:0.3 million (1:312,419) chance of being infected with COVID-19 in the airplane cabin.

Due to developing availability and reliability of COVID-19 testing and localized variability in reporting since the start of the pandemic, statistics for asymptomatic persons, infectiousness, and underreporting carry a degree of uncertainty which affects the overall risk calculation. The CDC indicates that asymptomatic cases could range from 10–70% of the infected population and that asymptomatic infectiousness ranges from 0.25–1.00 [12]. The Chow et al. study indicates a 95% credible interval for an estimated 10-fold underreporting factor ranges from 2.2–50 [14]. For this paper’s treatment of risk, uncertainty is established by first identifying the uncertainty in factors *f*_*a*_ and *f*_*c*_ in Equation 1, next fitting triangular distributions to each statistic with the peak at the baseline values stated above, then running 1 million simulations with a draw from each distribution to calculate risk, and finally selecting the 2.5% and 97.5% risk estimate from the 1 million simulations which set the bounds of the uncertainty interval. Using the above literature estimates of uncertainty in the parameters of *f*_*a*_ and *f*_*c*_, uncertainty in the transmission risk is estimated to range from 1 case for every 0.7 million (712,000) travelers to 1 case for every 8 million travelers.

At the onset of the pandemic in March, the estimated risk of transmission was 2.353×10^−6^ or 1:425,062 (*f*_*c*_=10). Estimated risk of being infected with COVID-19 could have been as high as 1:78,715 (1.27×10^−5^, *f*_*c*_=54). In comparison, estimated risk during the pandemic (April– September 2020) ranges from 1:1.3 million (*f*_*c*_=54) to 1:7.1 million (*f*_*c*_=10) with reduced passenger traffic and few documented secondary cases.

## 4. Discussion

Fewer than 50 passenger-related secondary cases have been recorded in the air travel system since the outbreak of COVID-19. There is documented evidence of flights with multiple infectious passengers but few or no secondary cases recorded [9] [19] [20]. Moreover, the majority of the documented secondary cases occurred on flights in March 2020 or earlier, when current common practices like wearing face masks and increased sanitization varied by region [21]. A notable finding of the Chen et al. [10] study discussed above is that only one possible secondary case was identified despite the large fraction of passengers on this flight, next to, in front of and behind index passengers. While physical distancing recommendations of many health organizations worldwide recommend 6 feet of separation to lower transmission risk [22], the results of this study indicate the actual risk of transmission in an aircraft cabin is still very low despite the relatively close proximity of passengers. This finding warrants a detailed analysis of the aircraft cabin environment that could explain how such results are possible [23].

The ICAO traveler population numbers, used in risk calculations presented herein, include global passengers, from regions such as Africa with less immediate outbreaks of COVID-19. Their inclusion could lower the ratio of infectious passengers to all passengers. Risk is also lowered by decreased worldwide air travel starting in March 2020 and limitation of air travelers to a small self-selecting subset of the global population, therefore a 10-fold change is assumed sufficient for a baseline estimated risk of transmission during air travel. Recognizing that studies could be ongoing to identify transmission events in the air travel system from summer 2020, the estimated transmission risk calculated for January–June 2020, 5.927×10^−7^ or 1:1.7 million (*f*_*c*_=10), should be considered the baseline estimated risk of transmission. With uncertainty, this risk ranges from 1 case for every 0.7 million (712,000) travelers to one case for every 8 million travelers.

While the risk of transmission in March 2020 in the general population was higher than current estimates, SARS-CoV-2 transmission aboard an airplane in March was still a low probability event. It should be noted that these calculations are based on limited testing of 2020 travelers and limited publicly available data, and are subject to the forces of a shifting global pandemic. While the specific numbers will change as more flights are made, tests performed, and papers published, the reports for dates covered by this study indicate a low risk of transmission aboard an aircraft. The inclusion of the two factors for asymptomatic transmission and underreporting are a novel approach to evaluating air travel risk for COVID-19 and lessen the impact of confounding factors and heterogeneous pandemic studies. This is a more nuanced approach to assessing COVID-19 air travel transmission risk than assuming a high degree of risk from select worst case scenarios; the probability of an infectious person aboard an airplane is not equal to the probability of a SARS-CoV-2 transmission to another person aboard that aircraft.

## 5. Conclusion

Data available during 2020 has shown that the risk of known communicable diseases being transmitted aboard aircraft is low [8]. This conclusion is consistent with studies on disease transmission in flight from other viruses such as from SARS-CoV-1 [5], Influenza [6], and Tuberculosis [7]. Similarly, the risk of SARS-CoV-2 transmission on an aircraft is also low, approximately 1:1.7 million. Given the global spread of COVID-19, it is clear that infectious individuals, including asymptomatic individuals, have been traveling by air. However, there has not been the large number of secondary cases that might be expected. Analysis of the limited reporting on infections traced to the air travel environment during the current COVID-19 pandemic supports the conclusion that even with infectious persons onboard, the overall risk of contracting COVID-19 from such an index passenger is low[9] [10] [19] [20], [24]-[29]. In addition, the improvement in safety provided by wearing masks is well established [30] [31] [32] and the results presented herein, particularly the reduction in estimated risk observed from April through September when masks were widely used in the air travel system, supports the recommendation that masks be worn properly at all times when in public spaces, including the aircraft cabin.

It is thought that the environmental control systems on the aircraft, which are more effective than those in common commercial spaces that the public may experience, contribute to the low probability of infection in the air travel system. Specifically, the high airflow, high-efficiency particulate air (HEPA) filtration, and seat-height/passenger-positioning (passengers do not face one another for the majority of the flight) are thought to minimize air flow between rows and protect passengers from infectious particle transfer. Studies of the engineering controls on aircraft are currently being prepared for publication by the authors.

## Supporting information

MOOSE Checklist

Boeing Assignment of Copyright 2021

## Data Availability

All inputs and outputs reported in this paper are provided in the main text. Researchers are encouraged to contact the corresponding author for clarifications, advice on data utilization, and collaborations.

## Acknowledgments

The authors wish to thank Dr. David Powell, Medical Advisor for IATA and Dr. Jason Armstrong, for thoughtful discussions on risk. We would also like to thank Dr. Armstrong for helping in the organization of this paper.

## Author contributions

J. K. Pang: Methodology, Investigation, Formal analysis, Writing - Original Draft, Writing - Review & Editing; S. P. Jones: Methodology, Formal analysis, Writing - Original Draft, Writing - Review & Editing; L. L. Waite: Methodology, Formal analysis; N. A. Olson: Conceptualization, Investigation, Writing – Review & Editing; R. J. Atmur: Conceptualization; J. J. Cummins: Conceptualization, Project administration, Writing - Review & Editing.

## References

[1] World Health Organization. Archived: WHO Timeline - COVID-19, https://www.who.int/news/item/27-04-2020-who-timeline---covid-19; 2020 [accessed 21 September 2020].

[2] World Health Organization. WHO Field Visit to Wuhan, China 20-21 January 2020, https://www.who.int/china/news/detail/22-01-2020-field-visit-wuhan-china-jan-2020; 2020 [accessed 21 September 2020].

[3] World Health Organization. Novel Coronavirus (2019-nCoV): Strategic Preparedness and Response Plan; Draft as of February 3, 2020. World Health Organization.https://www.who.int/docs/default-source/coronaviruse/srp-04022020.pdf; 2020 [accessed 21 September 2020].

[4] Ellwood M. Coronavirus Air Travel: These Numbers Show the Massive Impact of the Pandemic. Conde Nast Traveler. April 13, 2020 [accessed 21 September 2020]. www.cntraveler.com/story/coronavirus-air-travel-these-numbers-show-the-massive-impact-of-the-pandemic.

[5] Olsen SJ, Chang HL, Cheung TYY, Tang AFY, Fisk TL, Ooi SPL et al. Transmission of the Severe Acute Respiratory Syndrome on Aircraft. N Engl J Med 2003;349:2416–22. https://doi.org/10.1056/NEJMoa031349

[6] Leitmeyer K, Adlhoch C. Influenza Transmission on Aircraft, A Systematic Literature Review. Epidemiology 2016;27:743–51. https://doi.org/10.1097/EDE.0000000000000438

[7] World Health Organization. Tuberculosis on aircraft. In: World Health Organization. Tuberculosis and Air Travel: Guidelines for Prevention and Control. 3rd ed. World Health Organization; 2008: chap 2. Available from: https://www.ncbi.nlm.nih.gov/books/NBK143710/

[8] Mangili A, Gendreau MA. Transmission of infectious diseases during commercial air travel. Lancet 2005;365:989–96. https://doi.org/10.1016/S0140-6736(05)71089-8

[9] Bae SH, Shin H, Koo H-Y, Lee SW, Yang JM, Yon DK. Asymptomatic transmission of SARS-CoV-2 on evacuation flight. Emerg Infect Dis 2020;26:2705–8. https://doi.org/10.3201/eid2611.203353

[10] Chen J, He H, Cheng W, Liu Y, Sun Z, Chai C et al. Potential transmission of SARS-CoV-2 on a flight from Singapore to Hangzhou, China: An epidemiological investigation. Travel Med Infect Dis 2020;36:101816. https://doi.org/10.1016/j.tmaid.2020.101816

[11] Khanh NC, Thai PQ, Quach HL, Thi NAH, Dinh PC, Duong TN et al. Transmission of severe acute respiratory syndrome coronavirus 2 during long flight. Emerg Infect Dis 2020;26:2617–24. https://doi.org/10.3201/eid2611.203299

[12] United States Centers for Disease Control and Prevention. COVID-19 Pandemic Planning Scenarios. https://www.cdc.gov/coronavirus/2019-ncov/hcp/planning-scenarios.html; 2020 [accessed 8 October 2020].

[13] Rahmandad H, Lim TY, Sterman J. Estimating COVID-19 under-reporting across 86 nations: implications for projections and control. medRxiv. Preprint, version 2. Preprint posted online August 3, 2020. https://doi.org/10.1101/2020.06.24.20139451

[14] Chow CC, Chang JC, Gerkin RC, Vattikuti S. Global predication of unreported SARS-CoV2 infection from observed COVID-19 cases. medRxiv. Preprint posted online May 5, 2020. https://doi.org/10.1101/2020.04.29.20083485

[15] Hortaçsu A, Liu J, Schwieg T. Estimating the fraction of unreported infections in epidemics with a known epicenter: An application to COVID-19. J Econom. In press. Published online September 7, 2020. https://doi.org/10.1016/j.jeconom.2020.07.047

[16] Lau H, Khosrawipour T, Kobach P, Ichii H, Bania J, Khosrawipour V. Evaluating the massive underreporting and undertesting of COVID-19 cases in multiple global epicenters. Pulmonology. In press. Published online 6 June 2020. https://doi.org/10.1016/j.pulmoe.2020.05.015

[17] Effects of Novel Coronavirus (COVID-19) on Civil Aviation: Economic Impact Analysis. Air Transport Bureau, International Civil Aviation Organization. https://www.icao.int/sustainability/Documents/COVID-19/ICAO_Coronavirus_Econ_Impact.pdf; 2020 [accessed 13 October 2020].

[18] World Health Organization. Advice on use of masks in the context of COVID-19: interim guidance, 5 June 2020. World Health Organization. https://apps.who.int/iris/bitstream/handle/10665/332293/WHO-2019-nCov-IPC_Masks-2020.4-eng.pdf; 2020 [accessed 13 October 2020].

[19] Freedman DO, Wilder-Smith A. In-flight Transmission of SARS-CoV-2: a review of the attack rates and available data on the efficacy of face masks. J Travel Med. 2020;27:taaa178. https://doi.org/10.1093/jtm/taaa178

[20] Pavli A, Smeti P, Hadjianastasiou S, Theodoridou K, Spilioti A, Papadima K et al. In-flight transmission of COVID-19 on flights to Greece: An epidemiological analysis. Travel Med Infect Dis. 2020;38:101882. https://doi.org/10.1016/j.tmaid.2020.101882

[21] Elachola H, Ebrahim SH, Gozzer E. COVID-19: Facemask use prevalence in international airports in Asia, Europe and the Americas, March 2020. Travel Med Infect Dis. 2020;35:101637. https://doi.org/10.1016/j.tmaid.2020.101637

[22] United States Centers for Disease Control and Prevention. Social Distancing, Keep a Safe Distance to Slow the Spread. https://www.cdc.gov/coronavirus/2019-ncov/prevent-getting-sick/social-distancing.html; 2020 [accessed 21 September 2020].

[23] Silcott D, Kinahan S, Santarpia J, Silcott B, Silcott R, Silcott P et al. TRANSCOM/AMC Commercial Aircraft Cabin Aerosol Dispersion Tests. United States Transport Command, US Dept of Defense, https://www.ustranscom.mil/cmd/docs/TRANSCOM%20Report%20Final.pdf; 2020 [accessed 16 October 2020].

[24] Schwartz KL, Murti M, Finkelstein M, Leis JA, Fitzgerald-Husek A, Bourns L et al. Lack of COVID-19 transmission on an international flight. CMAJ. 2020;192:E410. https://doi.org/10.1503/cmaj.75015

[25] Eldin C, Lagier JC, Mailhe M, Gautret P. Probable aircraft transmission of Covid-19 inflight from the Central African Republic to France. Travel Med Infect Dis. 2020;35:101643. https://doi.org/10.1016/j.tmaid.2020.101643

[26] Hoehl S, Karaca O, Kohmer N, Westhaus S, Graf J, Goetsch U et al. Assessment of SARS-CoV-2 Transmission on an International Flight and Among a Tourist Group. JAMA Netw Open. 2020;3:e2018044. https://doi.org/10.1001/jamanetworkopen.2020.18044

[27] Choi EM, Chu DKW, Cheng PKC, Tsang DNC, Peiris M, Bausch DG et al. In-flight transmission of SARS-CoV-2. Emerg Infect Dis 2020;26:2713–6. https://doi.org/10.3201/eid2611.203254

[28] Speake H, Phillips A, Chong T, Sikazwe C, Levy A, Lang J et al. Flight-associated transmission of severe acute respiratory syndrome coronavirus 2 corroborated by whole-genome sequencing. Emerg Infect Dis 2020;26:2872–80. https://doi.org/10.3201/eid2612.203910

[29] Powell D. COVID-19 and borders, Considerations relating to travel and aviation. International Air Transport Association, https://www.iata.org/contentassets/f1163430bba94512a583eb6d6b24aa56/pha-webinar.pdf; 2020 [accessed 9 October 2020].

[30] Leung NHL, Chu DKW, Shiu EYC, Chan K-H, McDevitt JJ, Hau BJP et al. Respiratory virus shedding in exhaled breath and efficacy of face masks. Nat Med 2020;26:676–80. https://doi.org/10.1038/s41591-020-0843-2

[31] Loeb M, Dafoe N, Mahony J, John M, Sarabia A, Glavin V et al. Surgical mask vs N95 respirator for preventing influenza among health care workers, A randomized trial. JAMA 2009;302:1865–71. https://doi.org/10.1001/jama.2009.1466

[32] Bahl P, Bhattacharjee S, de Silva C, Chughtai AA, Doolan C, MacIntyre CR. Face coverings and mask to minimise droplet dispersion and aerosolisation: a video case study. Thorax 2020;75:1024–5. https://doi.org/10.1136/thoraxjnl-2020-215748

[33] International Air Transport Association. Restarting aviation following COVID-19. International Air Transport Association, www.iata.org/contentassets/f1163430bba94512a583eb6d6b24aa56/covid-medical-evidence-for-strategies-200504.pdf; 2020 [accessed 8 October 2020].

[34] Duncan I. Nearly 11,000 people have been exposed to the coronavirus on flights, the CDC says. The Washington Post. September 19, 2020 [accessed 21 September 2020]. www.washingtonpost.com/local/trafficandcommuting/nearly-11000-people-have-been-exposed-to-the-coronavirus-on-flights-the-cdc-says/2020/09/19/d609adbc-ed27-11ea-99a1-71343d03bc29_story.html.

