## Supplementary material for "Probability and Estimated Risk of SARS-CoV-2 Transmission in the Air Travel System: *A Systemic Review and Meta-Analysis*": MOOSE Checklist

**MOOSE Checklist for Meta-analyses of Observational Studies**

| **Item No** | **Recommendation** | **Reported on Page No** |
| --- | --- | --- |
| Reporting of background should include | | |
| 1 | Problem definition | 2 |
| 2 | Hypothesis statement | 2-3 |
| 3 | Description of study outcome(s) | 7-9, Table 1 |
| 4 | Type of exposure or intervention used | - |
| 5 | Type of study designs used | 3-7 |
| 6 | Study population | 4-6 |
| Reporting of search strategy should include | | |
| 7 | Qualifications of searchers (eg, librarians and investigators) | Title page, 4 |
| 8 | Search strategy, including time period included in the synthesis and key words | 4 |
| 9 | Effort to include all available studies, including contact with authors | 4-5 |
| 10 | Databases and registries searched | 4 |
| 11 | Search software used, name and version, including special features used (eg, explosion) | 4 |
| 12 | Use of hand searching (eg, reference lists of obtained articles) | Table 1 |
| 13 | List of citations located and those excluded, including justification | 5, Table 1 |
| 14 | Method of addressing articles published in languages other than English | - |
| 15 | Method of handling abstracts and unpublished studies | 4 |
| 16 | Description of any contact with authors | 5, 16 |
| Reporting of methods should include | | |
| 17 | Description of relevance or appropriateness of studies assembled for assessing the hypothesis to be tested | 4-5 |
| 18 | Rationale for the selection and coding of data (eg, sound clinical principles or convenience) | 3, 5-6 |
| 19 | Documentation of how data were classified and coded (eg, multiple raters, blinding and interrater reliability) | Table 1 |
| 20 | Assessment of confounding (eg, comparability of cases and controls in studies where appropriate) | 4 |
| 21 | Assessment of study quality, including blinding of quality assessors, stratification or regression on possible predictors of study results | 4-5 |
| 22 | Assessment of heterogeneity | 4 |
| 23 | Description of statistical methods (eg, complete description of fixed or random effects models, justification of whether the chosen models account for predictors of study results, dose-response models, or cumulative meta-analysis) in sufficient detail to be replicated | 6-7 |
| 24 | Provision of appropriate tables and graphics | 6, Table 1 |
| Reporting of results should include | | |
| 25 | Graphic summarizing individual study estimates and overall estimate | Table 2 |
| 26 | Table giving descriptive information for each study included | Table 1 |
| 27 | Results of sensitivity testing (eg, subgroup analysis) | Table 2, 11-12 |
| 28 | Indication of statistical uncertainty of findings | 11-12 |

| **Item No** | **Recommendation** | **Reported on Page No** |
| --- | --- | --- |
| Reporting of discussion should include | | |
| 29 | Quantitative assessment of bias (eg, publication bias) | 13-14 |
| 30 | Justification for exclusion (eg, exclusion of non-English language citations) | - |
| 31 | Assessment of quality of included studies | 7-10, Figure 1 |
| Reporting of conclusions should include | | |
| 32 | Consideration of alternative explanations for observed results | 13-15 |
| 33 | Generalization of the conclusions (ie, appropriate for the data presented and within the domain of the literature review) | 14-15 |
| 34 | Guidelines for future research | 14-15 |
| 35 | Disclosure of funding source | 16 |

*From*: Stroup DF, Berlin JA, Morton SC, et al, for the Meta-analysis Of Observational Studies in Epidemiology (MOOSE) Group. Meta-analysis of Observational Studies in Epidemiology. A Proposal for Reporting. *JAMA*. 2000;283(15):2008-2012. doi: 10.1001/jama.283.15.2008.
