## Supplementary material for "Probability and Estimated Risk of SARS-CoV-2 Transmission in the Air Travel System: *A Systemic Review and Meta-Analysis*": Boeing Assignment of Copyright 2021

**The Boeing Company**
5301 Bolsa Ave. H021-F241
Huntington Beach, Ca 92647

### ASSIGNMENT OF COPYRIGHT
### (“Agreement”)

Date of Publication: 4/9/2021

Publisher: MedRxiv (“Publisher(s)”)

Work Title: Probability and Relative Risk of SARS-CoV-2 Transmission on an Aircraft (“Work”)

Boeing Employee(s): Jenna K Pang

At least one, but possibly not all, of the authors of the Work is an employee of The Boeing Company or one of its wholly-owned subsidiaries (“BOEING”) who prepared the Work within the scope of his or her employment. The Boeing employee(s) listed above are not authorized to assign or license Boeing’s rights to the Work, or otherwise bind Boeing. However, subject to the limitations set forth below, Boeing is willing to assign its copyright in the Work to Publisher.

Notwithstanding any assignment or transfer to the Publisher, or any other terms of this Agreement, the rights granted by Boeing to Publisher are limited as follows: (i) any rights granted by Boeing to the Publisher are limited to the work-made-for-hire rights Boeing enjoys in the Work; (ii) Boeing makes no representation or warranty of any kind to the Publisher or any other person or entity regarding the Work, the information contained therein, or any related copyright; and (iii) Boeing retains a non-exclusive, perpetual, worldwide, royalty-free right, without restriction or limitation, to use, reproduce, publicly distribute, display, and perform and make derivative works from the Work, and to permit others to do so.

Brianna Kohlenberg
Contracts Specialist
Boeing Intellectual Property
